## Supplemental Table 1 for "Seroprevalence of SARS-CoV-2 and risk factors for infection among children in Uganda: a serial cross-sectional study"

| Vaccinated for COVID-19 | | 1,753 (52.7%) |
| --- | --- | --- |
| If vaccinated, number of doses | One dose  Two doses | 1,324 (75.4%)  431 (24.6%) |
| Type of vaccine (1^st^ dose) | Don’t know  Astra-Zeneca  Sinovac/Sinopharm  Johnson & Johnson  mRNA NOS  Moderna  Pfizer | 40 (2.3%)  709 (40.4%)  183 (10.4%)  518 (29.5%)  159 (9.1%)  87 (5.0%)  57 (3.3%) |
| Type of vaccine (2^nd^ dose) | Don’t know  Astra-Zeneca  Sinovac/Sinopharm  Johnson & Johnson  mRNA NOS  Moderna  Pfizer | 16 (3.7%)  258 (59.9%)  79 (18.3%)  4 (0.9%)  33 (7.7%)  20 (4.6%)  21 (4.9%) |

Supplemental Table 1. Vaccination status in 3,326 adults at follow-up survey.
